# Associations of Pathogenic Sequence Variants and Copy Number Variants with Cardiovascular and Major Clinical Outcomes in Adults with Congenital Heart Disease

**DOI:** 10.64898/2026.11.05.26367322

**Authors:** Peter Youssef, Florent Broussous, Haitham Alhussain, Karnaditya Rana, Mohammad Al-Najjar, Mohammad Modassir Islam, Alidjanov Xodjiakbar Kashipovich, Harshita Catherine Dasi, Mohammad Husnah Aman, Mansoor Ali Khan Kabul, Priya Chauhan

## Abstract

**Background:** Genetic abnormalities play a role in the phenotypic heterogeneity of congenital heart disease (CHD), but their associations with long-term cardiovascular outcomes in adults with congenital heart disease (ACHD) remain inadequately defined. This study examined the correlations between pathogenic sequence variations and copy number variants (CNVs) with cardiovascular and significant clinical outcomes in persons suffering from CHD.

**Methods:** This prospective observational cohort study included 1,000 adults with structurally confirmed CHD recruited from a tertiary care centre in Noida, India, between January 2022 and January 2026, with follow-up through August 2026. Those who have consented received exome sequencing and chromosomal microarray assessment and were categorized into four distinct, non-overlapping groups: reference, sequence-variant-only, CNV-only, and dual-positive. The primary outcome was the time to the first occurrence of any of the following events: cardiovascular death, unplanned heart failure hospitalization, resuscitated cardiac arrest, qualifying sustained ventricular tachyarrhythmia, heart transplantation or durable mechanical circulatory support. Appropriate statistical analysis was performed and results were obtained.

**Results:** The results showed that among the participants recruited, 800 were in the reference group, 120 had sequence variants alone, 60 had CNVs alone, and 20 had both sequence variants and CNVs. Over a median follow-up of 4.2 years, the primary outcome occurred in 12.0%, 20.0%, 25.0%, and 30.0% of participants, respectively. In the fully adjusted model, the sequence-variant-only group had an HR of 1.65 (95% CI, 1.01–2.69; nominal p = 0.0455), while the CNV-only group had an HR of 1.92 (95% CI, 1.04–3.52; nominal p = 0.0360), compared with the reference group. After Holm adjustment, neither association reached statistical significance (adjusted p = 0.072 for both comparisons). The dual-positive group had an HR of 2.12 (95% CI, 0.83–5.42; p = 0.1182). Extracardiac congenital anomalies and neurological disorders were more prevalent in those with genetic variations. Nonetheless, the secondary outcomes revealed no statistically significant correlations.

**Conclusions:** Pathogenic sequence variations and CNVs exhibited a numerically elevated adjusted risk for the primary cardiovascular composite outcome in ACHD, however neither primary association reached statistical significance after multiplicity adjustment. The findings highlight the potential significance of genetic diversity concerning clinical variability in ACHD, however they do not confirm independent prognostic significance.

## Introduction

Congenital heart disease (CHD) refers to structural abnormalities of the heart or great blood vessels that develop before birth and are present at birth, although they may be diagnosed later in life. Advances in cardiovascular diagnosis, surgical repair, and long-term care have improved survival among individuals with CHD, allowing an increasing number to reach adulthood [1]. However, adults with congenital heart disease (ACHD) continue to experience substantial morbidity and premature mortality. Their long-term outcomes vary according to cardiac anatomy, physiological status, and associated comorbidities [2]. Epidemiological studies also indicate that differences in survival persist into adulthood, highlighting the need to identify factors that may contribute to these variations in long-term outcomes [3,4].

Genetic variability is a key factor in the development and the phenotypic variability of CHD [5]. These include pathogenic sequence variants, such as single-nucleotide variants and small insertions or deletions, as well as copy number variants (CNVs), which involve the deletion or duplication of larger segments of DNA [6, 7, 8]. These modifications may influence genes and regulatory mechanisms pertinent to heart morphogenesis and the formation of additional organ systems [9, 10, 11]. Thus, clinical implications of a genetic diagnosis might involve not only the anatomical heart anomaly but also extracardiac symptoms and broader health risks. The existing ACHD recommendations acknowledge the importance of genetic assessment, especially for those with a familial background of CHD or clinical indicators of a genetic disorder [12, 13].

Earlier genomic studies have shown that the genetic factors underlying congenital heart disease (CHD) may also be associated with abnormalities in other organ systems. Homsy and colleagues studied 1,213 parent–offspring trios and found an increased burden of damaging de novo variants in genes associated with heart and brain development. These variants accounted for an estimated 20% of CHD cases accompanied by neurodevelopmental disabilities and other congenital anomalies, compared with around 2% of isolated CHD cases [14]. The findings suggest that cardiac and extracardiac abnormalities may share common genetic and developmental mechanisms, highlighting the need to examine the wider clinical effects of genetic variation.

Studies also indicate that genetic anomalies may correlate with prognosis. In the CHD GENES investigation, clinically relevant de novo variations were discovered in 11.7% of 2,517 individuals with CHD. In people with suitable surgical data, negative de novo mutations were associated with low transplant-free survival rate after the first open-heart surgery [15]. However, all these findings are mostly relevant to the postoperative period and cannot establish whether different types of genetic abnormalities are linked to heart diseases later in adult population.

Studies related to adults have demonstrated the predictive importance of specific genetic diseases. The 22q11.2 deletion syndrome is associated with increased mortality rates in individuals with tetralogy of Fallot. [16]. Recently, 9,826 ACHD patients demonstrated a correlation between clinically documented syndromic illnesses and overall mortality [17]. These findings establish that syndromic conditions are relevant to prognosis in ACHD. Nonetheless, syndromic diagnoses from clinical documentation do not correspond to exposure categories established from molecular identification of sequence variants and copy number variations. The two approaches analyze related but distinct challenges. One examines outcomes associated with recognized clinical syndromes, while the other evaluates correlations based on genetic variant categorizations, including findings that may not correspond directly with a known syndromic diagnosis [18].

Consequently, an important question to be addressed is whether molecularly characterized sequence variations and CNVs correlate with subsequent cardiovascular outcomes in ACHD, and whether these correlations persist after adjusting for demographic and clinical factors. Assessing these variant categories independently may elucidate whether their correlations with outcomes vary. This differentiation is significant as genetic anomalies differ in their biological impacts and phenotypic manifestations, and results from individual syndromes or restricted anatomical categories may not be universally applicable to the broader ACHD population.

The present study examined the associations between qualifying pathogenic or likely pathogenic sequence variants and CNVs and cardiovascular outcomes in adults with CHD. These associations were assessed before and after adjustment for prespecified demographic and clinical factors. We additionally investigated the impact of our results on genetic exposure categories to provide further details on the association between genetic variability and future cardiovascular events in ACHD.

## Methods

### Study design and setting

This was a prospective observational cohort study conducted at a tertiary care centre in Noida, Uttar Pradesh, India, to investigate the associations of pathogenic sequence variants and CNVs with cardiovascular and major clinical outcomes in ACHD. Participants were recruited consecutively from the specialist ACHD service, Noida between January 2022 and January 2026, with follow-up continuing until August 2026.

Baseline characteristics were established through participant assessment and review of clinical records available at enrolment. Enrolment marked the beginning of prospective outcome observation. Follow-up was conducted through hospital visits and electronic medical records. No experimental intervention or random allocation was undertaken, and clinical management remained under the responsibility of the treating teams.

### Sample Size

The sample-size was calculated using the non-central chi-square method implemented in G*Power version 3.1.9.7. Four mutually exclusive exposure groups were considered: reference, pathogenic sequence variant only, copy-number variant only, and dual-positive. Assuming a small effect size (Cohen’s w = 0.10), a significance level (α) of 0.05, statistical power of 80%, and 3 degrees of freedom, the minimum required sample size was 895 participants. After applying a 10% adjustment for anticipated data loss, the final sample size was estimated at 995 participants.

### Participants

Adults with structural CHD who were 18 years of age or older and whose condition was verified by an expert evaluation of cardiac imaging, surgical records, or other conclusive laboratory diagnosis were eligible. Participation, genetic testing, follow-up, and access to medical information were taken after obtaining informed consent. Duplicate enrolments, people with acquired cardiovascular illness without structural CHD, and people for whom valid permission could not be obtained through the authorized procedure were excluded. The exclusion criteria did not include recognized genetic syndromes, prior cardiovascular events, severe CHD, or neurodevelopmental disability on their own. When necessary, consent support and appropriate communication were given. Eligibility, recruitment, non-participation reasons, and inclusion in the analytical populations were all recorded in screening records.

### Genetic assessment and variant classification

All participants underwent a standardized core genetic assessment irrespective of syndromic appearance, disease severity or previous genetic testing. Peripheral venous blood samples were collected at the time of enrolment in EDTA tubes, and genomic DNA was extracted using the QIAamp DNA Blood Mini Kit (QIAGEN). DNA concentration was measured using the Qubit dsDNA HS Assay (Thermo Fisher Scientific), with sample acceptance based on the laboratory’s validated quality criteria. Exome libraries were prepared using the SureSelect Human All Exon V8 capture kit (Agilent Technologies) and sequenced on the NovaSeq 6000 platform (Illumina). Sequence reads were aligned to the GRCh38 human reference genome, and germline single-nucleotide variants and small insertions/deletions were identified using GATK HaplotypeCaller. Analysis focused on a prespecified set of genes associated with CHD and related syndromes. Chromosomal microarray analysis was performed using the CytoScan HD Array (Thermo Fisher Scientific), and copy-number changes were analysed using Chromosome Analysis Suite software. Sequence variants were classified according to ACMG/AMP criteria [19], while CNVs were interpreted using ACMG/ClinGen standards. Classification considered gene–disease validity, inheritance pattern, zygosity and disease mechanism. Qualifying pathogenic findings defined exposure-positive status; variants of uncertain significance did not independently qualify.

### Genetic exposure definitions

The primary exposures consisted of qualifying pathogenic sequence variants and CNVs linked to CHD or related syndromic disorders. Participants were classified into four distinct and non-overlapping groups. The sequence-variant-exclusive category consisted of participants with a qualifying sequence variant without a qualifying CNV. The CNV-only subgroup comprised of individuals exhibiting a qualifying CNV but lacking a qualifying sequence variant. The dual-positive group included individuals exhibiting both variant categories. The reference group consisted of individuals with no qualifying pathogenic findings detected by the study assays.

Variants of uncertain significance (VUS) were not considered pathogenic and were insufficient on their own to classify participants as genetically exposed. Their frequency was documented, and sensitivity analyses were performed to assess the effect of excluding these variants from the reference group.

### Baseline clinical assessment

Baseline data was obtained using standardized case-report forms through participant interviews, clinical assessments, cardiac imaging, and evaluation of medical records. Documented traits included congenital heart conditions, anatomical complexities, physiological status, New York Heart Association functional class, oxygen saturation levels, systemic ventricular function, residual lesions, prior corrective or palliative interventions, age at initial surgery, arrhythmias, past cardiovascular incidents, medications and conventional cardiovascular risk determinants. Extracardiac congenital anomalies, neurodevelopmental impairments, familial medical history, and related health access were also recorded.

Anatomical complexity was categorized as simple, moderate, or great complexity using the 2025 ACC/AHA ACHD classification. Classification was applied to baseline clinical information.

Clinical outcomes were ascertained through hospital visits and review of available clinical records, including hospital records, discharge summaries, procedure reports, imaging records, and documented mortality information. The dates of qualifying events and the end of follow-up for each participant were documented. Follow-up ended at the first occurrence of the primary outcome or at the end of the participant’s available observation period, including administrative censoring, as applicable. The ascertainment of outcomes was based on the available clinical documentation.

### Outcome Definitions and Ascertainment

The primary outcome was time from enrolment to the first occurrence of cardiovascular death, unplanned hospitalisation for heart failure, resuscitated cardiac arrest or sustained ventricular tachyarrhythmia requiring urgent treatment, or heart transplantation or durable mechanical circulatory support. Only the first qualifying event recorded for each participant was considered in the primary composite outcome analysis.

Cardiovascular death was defined as death attributable to a cardiovascular cause, as documented in mortality records or clinical records. It included sudden cardiac death, myocardial infarction, progressive heart failure, fatal cardiac arrhythmia or complications directly related to the underlying congenital cardiac defect. Unscheduled hospital admissions for heart failure were identified from medical records and discharge summaries that signified hospitalization due to worsened heart failure. Instances of resuscitated cardiac arrest and prolonged ventricular tachyarrhythmias requiring immediate intervention were identified from recorded clinical events and hospital documentation. Heart transplantation and long-term mechanical circulatory support were identified from procedural documentation.

Secondary outcomes included all-cause mortality, stroke or systemic embolism, atrial arrhythmia requiring hospitalization or intervention, unanticipated cardiovascular hospitalization, unexpected cardiac reintervention, and significant non-cardiovascular hospitalization. Each components of the primary composite was also assessed individually.

Event dates and the end of each participant’s observed follow-up were recorded. Outcome events were assessed by reviewers blinded to genetic exposure status to minimise bias. Only the first qualifying event contributed to the primary composite analysis.

### Follow-up

Follow-up duration was calculated from enrolment to the first qualifying outcome event, non-cardiovascular death, withdrawal, last confirmed outcome assessment, or administrative cutoff, as applicable. Participants without the outcome were censored at their last documented follow-up or the administrative study cutoff, whichever occurred first. Total observed person-time was calculated by summing individual follow-up durations.

Observation continued after nonfatal primary events to ascertain subsequent secondary outcomes. For all-cause mortality, follow-up continued until death, withdrawal, last confirmed vital-status assessment, or administrative cutoff, whichever occurred first.

Loss to follow-up was defined as the absence of further clinical follow-up or outcome information. Follow-up was conducted through hospital visits and review of electronic medical records. All 1,000 participants were retained until the end of the study, and no loss to follow-up was recorded.

In the cause-specific Cox analysis, non-cardiovascular deaths were treated as censoring events at the time of occurrence. Non-cardiovascular deaths were considered as a competing event when estimating cumulative incidence of cardiac death.

### Statistical Analysis

Data were analysed using SPSS version 26.0 and the baseline characteristics were summarized as mean ± standard deviation, median (interquartile range), or frequency and percentage using descriptive statistics. Cox regression estimated hazard ratios with 95% confidence intervals were adjusting and proportional hazards assumptions were assessed. Holm’s correction was applied to the two primary comparisons. Fine–Gray models accounted for competing death in selected nonfatal outcomes. Sensitivity analyses were done which assessed robustness, while dual-positive estimates were interpreted cautiously because of the small samples recruited. Two-sided p < 0.05 has indicated statistical significance; secondary and sensitivity analyses used unadjusted p-values.

### Ethical approval and informed consent

The study received approval from the Institutional Ethical Committee Board of the tertiary care center in Noida, Uttar Pradesh with number AUUP/78JAN/2022-4C. Informed consent was obtained from participants. Consent addressed sample storage, family implications, return of findings, future research and controlled data sharing. Participants received coded identifiers, and identifiable clinical and genomic information was stored securely with restricted access. Pre-test information and post-test counselling were provided. Potential risks associated with genetic testing and blood sampling were explained during the consent process.

## RESULTS

Of the 1,200 individuals assessed for eligibility, 150 were excluded for not meeting the eligibility criteria, while 50 declined participation. The final cohort comprised 1,000 participants, all of whom had interpretable sequence-variant and copy-number variant (CNV) assessment results. Based on the genetic assessment findings, participants were classified into four groups: reference (n = 800; 80.0%), sequence-variant-only (n = 120; 12.0%), CNV-only (n = 60; 6.0%), and dual-positive (n = 20; 2.0%). (Figure 1)

**Figure 1.**
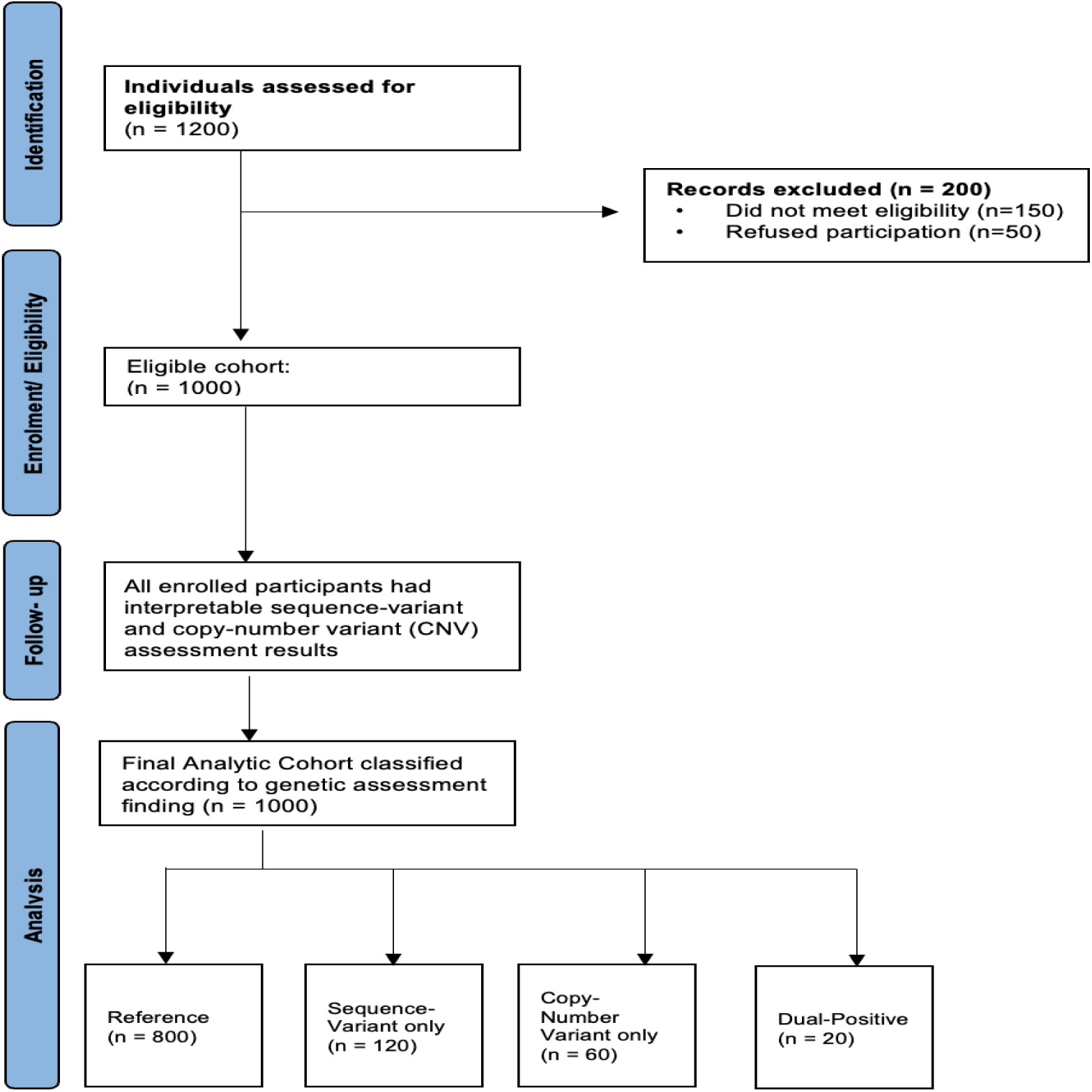
STROBE Flow Diagram of Participant Enrolment, Eligibility Assessment, and Genetic Classification. Mean age was lower in the genetic exposure groups than in the reference group. Extracardiac congenital anomalies (8.0%, 25.0%, 40.0%, and 45.0%, respectively) and neurodevelopmental disability (5.0%, 15.0%, 35.0%, and 40.0%, respectively) showed progressively higher frequencies across the four groups. Greater anatomical complexity, ventricular dysfunction, and previous heart-failure hospitalization were also more frequent in the CNV-only and dual-positive groups. Absolute standardized differences for extracardiac congenital anomalies versus the reference group were 0.471, 0.808, and 0.923 in the sequence-variant-only, CNV-only, and dual-positive groups, respectively. (Table 1)

**Table 1.** Baseline demographic and clinical characteristics by genetic exposure group.

| **Characteristic** | **Reference<br>(n = 800)** | **Sequence only<br>(n = 120)** | **CNV only<br>(n = 60)** | **Dual-positive<br>(n = 20)** |
|---|---|---|---|---|
| Age, years, mean ± SD | 35.8 ± 12.4 | 33.6 ± 11.2 | 31.9 ± 10.8 | 30.7 ± 9.6 |
| Female sex, n (%) | 416 (52.0) | 64 (53.3) | 29 (48.3) | 11 (55.0) |
| Simple anatomical complexity, n (%) | 240 (30.0) | 30 (25.0) | 9 (15.0) | 3 (15.0) |
| Moderate anatomical complexity, n (%) | 400 (50.0) | 66 (55.0) | 36 (60.0) | 11 (55.0) |
| Great anatomical complexity, n (%) | 160 (20.0) | 24 (20.0) | 15 (25.0) | 6 (30.0) |
| Previous corrective/palliative surgery, n (%) | 624 (78.0) | 98 (81.7) | 51 (85.0) | 17 (85.0) |
| Age at first operation, years, median (Q1–Q3) | 3.0 (0.8–8.0) | 2.5 (0.6–7.0) | 1.8 (0.4–5.5) | 1.5 (0.3–5.0) |
| NYHA functional class III–IV, n (%) | 72 (9.0) | 14 (11.7) | 9 (15.0) | 3 (15.0) |
| Systemic ventricular dysfunction, n (%) | 112 (14.0) | 20 (16.7) | 10 (16.7) | 4 (20.0) |
| Oxygen saturation, %, mean ± SD | 96.2 ± 3.4 | 95.8 ± 3.8 | 95.1 ± 4.5 | 94.8 ± 4.7 |
| Previous heart-failure hospitalization, n (%) | 64 (8.0) | 12 (10.0) | 7 (11.7) | 3 (15.0) |
| Previous sustained ventricular arrhythmia, n (%) | 40 (5.0) | 8 (6.7) | 4 (6.7) | 1 (5.0) |
| Previous stroke/systemic embolism, n (%) | 24 (3.0) | 4 (3.3) | 2 (3.3) | 1 (5.0) |
| Extracardiac congenital anomaly, n (%) | 64 (8.0) | 30 (25.0) | 24 (40.0) | 9 (45.0) |
| Neurodevelopmental disability, n (%) | 40 (5.0) | 18 (15.0) | 21 (35.0) | 8 (40.0) |
| Hypertension, n (%) | 112 (14.0) | 15 (12.5) | 7 (11.7) | 2 (10.0) |
| Diabetes mellitus, n (%) | 32 (4.0) | 4 (3.3) | 3 (5.0) | 1 (5.0) |
| Current smoking, n (%) | 96 (12.0) | 12 (10.0) | 5 (8.3) | 2 (10.0) |

Values are presented as mean ± SD, median (Q1–Q3), or n (%). Percentages use non-missing denominators. Age at first operation was calculated among participants with previous cardiac surgery and a documented age at the first operation. Anatomical complexity and systemic ventricular dysfunction were classified using prespecified study criteria. CNV, copy-number variant; NYHA, New York Heart Association; SD, standard deviation.

The median follow-up duration was 4.2 years (Q1–Q3, 3.8–4.5), accounting for 4,284 total follow-up person-years. All 1,000 participants were retained for follow-up, with no loss to follow-up.

The primary composite outcome occurred in 12.0% of the reference group, compared with 20.0% of the sequence-variant-only group, 25.0% of the CNV-only group, and 30.0% of the dual-positive group. Cardiovascular death, unplanned heart-failure hospitalization, and resuscitated cardiac arrest or qualifying sustained ventricular tachyarrhythmia were also more frequent across the genetic exposure groups than in the reference group. Heart transplantation or durable mechanical circulatory support occurred in 1.0%, 2.5%, 3.3%, and 5.0% of participants, respectively. Overall, the CNV-only and dual-positive groups had higher crude proportions of adverse clinical outcomes. However, these comparisons do not account for differences in follow-up duration or indicate whether the observed differences are statistically significant (Table 2).

**Table 2.** Primary Clinical Outcomes According to Genetic Exposure Group During Follow-up.

| **Outcome** | **Reference**<br>**(n = 800)** | **Sequence only**<br>**(n = 120)** | **CNV only**<br>**(n = 60)** | **Dual-positive**<br>**(n = 20)** |
|---|---|---|---|---|
| Primary composite: first qualifying event | 96 (12.0) | 24 (20.0) | 15 (25.0) | 6 (30.0) |
| Cardiovascular death | 24 (3.0) | 6 (5.0) | 4 (6.7) | 2 (10.0) |
| Unplanned heart-failure hospitalization | 64 (8.0) | 16 (13.3) | 10 (16.7) | 4 (20.0) |
| Resuscitated cardiac arrest or qualifying sustained ventricular tachyarrhythmia | 24 (3.0) | 6 (5.0) | 4 (6.7) | 2 (10.0) |
| Heart transplantation or durable mechanical circulatory support | 8 (1.0) | 3 (2.5) | 2 (3.3) | 1 (5.0) |

Values are n (%). The primary composite counts each participant once at the first qualifying event; component outcomes may overlap. All-cause mortality includes cardiovascular deaths. Percentages represent crude event proportions over observed follow-up and are not cumulative-incidence estimates at a common time point.

All-cause mortality occurred in 42 (5.3%), 9 (7.5%), 6 (10.0%), and 3 (15.0%) participants in the reference, sequence-only, CNV-only, and dual-positive groups, respectively. Unplanned cardiovascular hospitalization was reported in 144 (18.0%), 30 (25.0%), 18 (30.0%), and 7 (35.0%) participants, while major non-cardiovascular hospitalization occurred in 120 (15.0%), 24 (20.0%), 15 (25.0%), and 6 (30.0%), respectively. These descriptive proportions do not establish independent associations between genetic exposure and clinical outcomes (Table 3).

**Table 3.** Secondary clinical outcomes during follow up by genetic exposure group.

| **Outcome** | **Reference (n = 800)** | **Sequence only (n = 120)** | **CNV only (n = 60)** | **Dual-positive (n = 20)** |
|---|---|---|---|---|
| All-cause mortality | 42 (5.3) | 9 (7.5) | 6 (10.0) | 3 (15.0) |
| Stroke/systemic embolism | 24 (3.0) | 5 (4.2) | 3 (5.0) | 1 (5.0) |
| Atrial arrhythmia requiring hospitalization/intervention | 80 (10.0) | 15 (12.5) | 9 (15.0) | 3 (15.0) |
| Unplanned cardiovascular hospitalization | 144 (18.0) | 30 (25.0) | 18 (30.0) | 7 (35.0) |
| Unplanned cardiac reintervention | 64 (8.0) | 10 (8.3) | 7 (11.7) | 2 (10.0) |
| Major noncardiovascular hospitalization | 120 (15.0) | 24 (20.0) | 15 (25.0) | 6 (30.0) |

Values are n (%). Participants with at least one event are reported for each outcome; categories may overlap. All-cause mortality includes cardiovascular deaths. Percentages represent crude event proportions over observed follow-up and are not cumulative-incidence estimates at a common time point.
CNV, copy-number variant.

In the fully adjusted model, the sequence-variant-only and CNV-only groups had higher hazard estimates for the primary composite outcome than the reference group (HR, 1.65; 95% CI, 1.01– 2.69; and HR, 1.92; 95% CI, 1.04–3.52, respectively). Although the nominal p-values were 0.0455 and 0.0360, respectively, neither association retained statistical significance after Holm adjustment (adjusted p = 0.0720 for both). The dual-positive group also showed a higher hazard estimate, with considerable uncertainty (HR, 2.12; 95% CI, 0.83–5.42; p = 0.1182). For secondary outcomes, the adjusted estimates were generally above 1, but their confidence intervals were wide and included the null value, providing limited evidence of associations with all-cause mortality, stroke/systemic embolism, or first unplanned cardiovascular hospitalization (Table 4).

**Table 4.** Associations of genetic exposure with primary and selected secondary outcomes.

| Outcome and model | Sequence only vs reference | CNV only vs reference | Dual-positive vs reference |
|---|---|---|---|
| Primary composite: unadjusted Cox model | 1.77 (1.13–2.78) | 2.32 (1.34–4.01) | 2.94 (1.29–6.71) |
| Primary composite: adjusted Model 1 | 1.75 (1.09–2.80) | 2.12 (1.18–3.81) | 2.46 (1.00–6.06) |
| Primary composite: adjusted Model 2 | 1.65 (1.01–2.69)<br>p = 0.0455 | 1.92 (1.04–3.52)<br>p = 0.0360 | 2.12 (0.83–5.42)<br>p = 0.1182 |
| All-cause mortality: adjusted Cox model | 1.38 (0.63–3.02)<br>p = 0.4237 | 1.73 (0.70–4.27)<br>p = 0.2318 | 2.10 (0.65–6.80)<br>p = 0.215 |
| Stroke/systemic embolism: adjusted Fine–Gray model | 1.22 (0.36–4.12)<br>p = 0.7470 | 1.49 (0.39–5.66)<br>p = 0.5564 | 1.80 (0.20–16.20)<br>p = 0.594 |
| First unplanned cardiovascular hospitalization: adjusted Fine–Gray model | 1.42 (0.92–2.18)<br>p = 0.1116 | 1.62 (0.93–2.80)<br>p = 0.0865 | 1.85 (0.85–4.05)<br>p = 0.122 |

Values are HRs or sHRs with 95% CIs. Model 1 adjusted for age, sex, and two ancestry principal components. Model 2 additionally adjusted for anatomical complexity, NYHA class, systemic ventricular dysfunction, previous cardiac surgery, heart-failure hospitalization, sustained ventricular arrhythmia, and stroke/systemic embolism. Non-outcome death was treated as a competing event in Fine–Gray models. P-values are two-sided; Holm adjustment was applied to the two primary Model 2 comparisons. Secondary-outcome p-values and 95% CIs were unadjusted for multiplicity. HR, hazard ratio; sHR, subdistribution hazard ratio; CI, confidence interval; NYHA, New York Heart Association.

Sensitivity analyses generally showed higher hazard estimates for the primary cardiovascular outcome in the sequence-variant-only and CNV-only groups than in the reference group (Table 5). In analyses restricted to pathogenic findings, the aHRs were 1.60 (95% CI, 0.92–2.77) for the sequence-variant-only group and 1.85 (95% CI, 0.91–3.75) for the CNV-only group. Excluding VUS carriers from the reference group yielded aHRs of 1.67 (95% CI, 1.004–2.78) and 1.95 (95% CI, 1.04–3.65), respectively. Among unrelated participants, the comparable HRs were 1.62 (95% CI, 0.97–2.70) for the sequence-variant-only group and 1.88 (95% CI, 1.00–3.52) for the CNV-only group. After removing recurrent 22q11.2 deletion carriers, the estimates were 1.58 (95% CI, 0.91–2.75) and 1.58 (95% CI, 0.68–3.67), respectively. The CIs were large in several analyses, particularly after removing recurrent 22q11.2 deletion carriers. Although the estimates were consistent in direction across sensitivity analyses, their magnitude and precision varied.

**Table 5.** Sensitivity analyses of genetic exposure and the primary cardiovascular outcome.

| Analysis population | Participants / primary events | Sequence only vs reference<br>aHR (95% CI) | CNV only vs reference<br>aHR (95% CI) |
|---|---|---|---|
| Pathogenic findings only | 960 / 130 | 1.60 (0.92–2.77)<br>p = 0.0932 | 1.85 (0.91–3.75)<br>p = 0.0875 |
| Excluding VUS carriers from reference | 920 / 133 | 1.67 (1.004–2.78)<br>p = 0.0486 | 1.95 (1.04–3.65)<br>p = 0.0369 |
| Excluding recurrent 22q11.2 deletions | 966 / 131 | 1.58 (0.91–2.75)<br>p = 0.105 | 1.58 (0.68–3.67)<br>p = 0.2874 |
| Unrelated participants only | 974 / 137 | 1.62 (0.97–2.70)<br>p = 0.0635 | 1.88 (1.00–3.52)<br>p = 0.0485 |
| Complete-case analysis | 1,000 / 141 | 1.65 (1.01–2.69)<br>p = 0.0455 | 1.92 (1.04–3.52)<br>p = 0.0360 |

Values are adjusted hazard ratios (aHRs) with 95% confidence intervals (CIs). P-values are nominal and unadjusted for multiple comparisons. Participant and event counts represent the retained cohort for each analysis. VUS, variant of uncertain significance.

## Discussion

The results of the study demonstrated numerical differences in adverse clinical outcomes across genetic exposure groups in ACHD population. Both the sequence-variant-only and CNV-only groups had higher crude event proportions and adjusted hazard ratios than the reference group, although neither primary comparison reached the prespecified level of statistical significance. The dual-positive group had the highest crude event proportion, but its small sample size made the estimate less precise. These findings suggest a possible relationship between genetic variation, multisystem manifestations and cardiovascular outcomes. However, they do not establish an independent prognostic effect or provide sufficient evidence to support changes in clinical management.

At baseline, those with qualifying genetic results were younger and exhibited greater frequencies of extracardiac congenital anomalies and neurodevelopmental disabilities compared to the reference group. This pattern was similar with the findings of Homsy et al., who discovered a surplus of deleterious de novo variations in genes essential in the developing heart and brain [12]. These finding are consistent with Zaidi et al., who identified damaging de novo mutations in histone-modifying genes in CHD [20]. The findings supported common developmental factors influencing both cardiac and extracardiac anomalies, although it did not confirm that genetic exposure was responsible for the baseline differences reported in this study [21]. Unplanned hospital admissions due to heart failure were the predominant primary outcome among all groups. Cardiovascular mortality, cases of revived cardiac arrest or qualifying ventricular tachyarrhythmia, and events of transplantation or prolonged mechanical circulatory support were also numerically more frequent among individuals with qualifying genetic results. Similar results were also reported by Watkins et al. [22]. Boskovski et al. reported reduced transplant-free survival rates postoperatively in patients with clinically significant, de novo alterations [15]. However, their analysis of combined sequence-variant and CNV exposure, along with their surgical group, differed from the present data, limiting direct comparison.

Adult studies provided further evidence concerning specific genetic diagnoses. Van Mil et al. and Kauw et al. reported poorer survival among adults with 22q11.2 deletion syndrome and selected congenital cardiac diagnoses [16,23]. These findings supported the potential long-term relevance of a molecular diagnosis but could not be generalized to all pathogenic CNVs. Furthermore, mortality findings from these studies were not directly comparable with the present composite outcome, which included nonfatal hospitalizations and interventions.

The associations between genetic findings and adverse cardiovascular outcomes became weaker after adjustment for demographic and clinical characteristics. This suggested that differences in the participants’ clinical profiles may have partly contributed to the observed associations. However, the reduction in the estimates did not establish how much of the association was explained by these factors. Anatomical complexity, ventricular dysfunction and previous cardiac events may themselves have been consequences of the underlying genetic condition [8,24]. Adjusting for these characteristics could therefore have accounted for part of the pathway through which genetic findings influenced later outcomes. The final model represented the association remaining after these clinical factors were considered, rather than the overall influence of the genetic condition. After adjustment for multiple comparisons, the primary associations did not meet the prespecified threshold for statistical significance. Although the confidence intervals excluded the null value, they were not adjusted for multiplicity, whereas the primary p-values accounted for multiple testing. The findings therefore remained compatible with potentially important associations, but their magnitude and reproducibility were uncertain.

Although the CNV-only group had a higher hazard estimate, a stronger association relative to the sequence-variant-only group could not be established because the two groups were not compared directly in a statistical analysis.

The findings for the dual-positive group warranted particular caution because the small number of participants and outcome events limited the precision of the estimates. Despite the group’s demonstration of an increased crude ratio of poor outcomes, the available data were inadequate to estimate the degree of the association. The coexistence of sequence variations and CNVs did not demonstrate any additional or synergistic effects. Previous studies have highlighted the contribution of recurrent CNVs to CHD, emphasizing their variable penetrance, pleiotropic effects and potential interactions with additional genetic variant [25]. Extensive exposure categories may have concealed differences in disease mechanisms, penetrance, and clinical outcomes.

The secondary outcomes exhibited a fairly similar descriptive pattern. Participants with qualifying genetic results frequently exhibited a higher frequency of all-cause death, unexpected cardiovascular hospitalizations, and significant non-cardiovascular hospitalizations. The crude proportions of atrial arrhythmias requiring hospitalization or intervention were numerically higher across the genetic exposure groups. Stroke or systemic embolism remained relatively uncommon, while cardiac reintervention showed no consistent numerical pattern across the groups. This was consistent with Bouchardy et al., who reported that cardiovascular complications, including heart failure and arrhythmia, remain important contributors to morbidity and mortality among ACHD [26]. The greater frequency of non-cardiovascular hospitalization may have reflected associated extracardiac conditions as also stated by Singh et al. [27]. However, the available data did not permit confirmation of this explanation. Differences in follow-up practices, access to healthcare and clinical judgement may have also affected hospitalization and intervention rates as also supported by Benderly et al. [28]. Therefore, the observed differences in secondary outcomes cannot be attributed to genetic findings alone. After adjustment for relevant factors, no statistically significant associations were found between genetic exposure and mortality, stroke or systemic embolism, or unplanned cardiovascular hospitalization.

The estimates had limited precision, and the absence of statistically significant findings did not indicate that outcomes were equivalent between groups. Mortality hazard ratios and competing-risk estimates also needed to be interpreted separately, as they address different aspects of risk. Ghimire et al. reported greater healthcare resource use among infants with 22q11.2 deletion but found no significant association with adjusted in-hospital mortality [29]. Differences in the study populations and outcomes measured should be considered when interpreting these findings. Sensitivity analyses produced modest changes following alternative exposure definitions, exclusion of uncertain-variant carriers and restriction to unrelated participants. The findings did not show that classification uncertainty or familial clustering accounted for the observed associations. Nominally significant results in selected analyses did not alter the conclusions from the multiplicity-adjusted primary analyses. Upon the exclusion of recurrent 22q11.2 deletion carriers, the hazard estimate for the CNV-only group decreased although the confidence interval indicated increased ambiguity. While this finding corresponds to previous studies on the deletion, it may indicate alterations in case composition or diminished statistical accuracy instead of a direct consequence of the deletion [16, 23, 30, 31]. The sensitivity analyzes included overlapping samples and hence did not represent independent validation. Likewise, the complete-case analysis did not provide any further evidence of robustness since its results were unchanged.

The genetic evaluation may have clinical relevance even if the role of genetic exposure categories as a prognostic factor is not clear. The significance of coordination between ACHD and genetics was brought out by Oehlman et al. without showing an improvement in the cardiovascular outcome [13]. In accordance with current ACHD recommendations [14], the findings reported do not recommend modifications in therapy or monitoring purely based on genetic exposure category. Clinical decisions must to take into account the precise genetic diagnosis in conjunction with cardiac anatomy, physiology, and related extracardiac diseases.

A few limitations of this study should be acknowledged. The precision of the results was reduced by limited sample sizes of exposed individuals and infrequent results, although overly generalized genetic categories may have contributed to the ambiguity between specific diagnoses. Recruitment from specialist services could limit generalizability, and an assay-negative result did not exclude genetic susceptibility. Crude event proportions did not represent risks over a fixed period. Future studies should include larger, independent ACHD cohorts with standardized clinical assessments, reliable genetic testing and reproducible analyses. Research should examine specific genetic diagnoses, account for competing risks and distinguish overall associations from those remaining after clinical adjustment. External validation would be needed to determine whether genetic information improved prediction and informed patient care. Future studies could assess whether automated ECG-based arrhythmia classification improves the consistency of rhythm outcome measurement in ACHD cohorts [32].

## Conclusion

In this population of patients with ACHD, the subgroups having only sequence variant and only CNV exhibited adjusted hazard estimates in relation to the major cardiovascular composite than the reference subgroup. Nonetheless, neither association retained statistical significance following the Holm adjustment, while the estimate for the dual-positive subgroup was ambiguous. The previously described phenotypes and sensitivity analysis underline the significance of variability in clinical and genetic categorization when assessing such associations. Further validation in independent groups is essential to determine whether genome-derived insights offer any clinical significance beyond clinical parameters.

## Transparency Statement

The corresponding author affirms that this manuscript provides an honest, accurate and transparent account of the study; no important aspects of the study have been omitted; and any deviations from the originally planned study have been appropriately disclosed and explained.

## Data Availability

The data that supports the findings of this study are available on request from the corresponding author. The data is not publicly available due to privacy or ethical restrictions.

## Data Availability

The data underlying this study are available from the corresponding author upon reasonable request. Individual-level genetic and clinical data are not publicly available because of participant privacy considerations and institutional ethics restrictions.

